# Inclusion of medical fitness to drive in medical postgraduate training curricula

**DOI:** 10.1101/2020.04.22.20075465

**Authors:** Laith Al Azawi, Aisling O’Byrne, Lily Roche, Desmond O’Neill, Margaret Ryan

## Abstract

**Background:** Transport mobility, and access to driving, is an important factor in social inclusion and well-being. Doctors have an important role to play in supporting safe mobility through applying the knowledge developed in the field of traffic medicine and incorporating state of the art national and international medical fitness to drive (MFTD) guidelines. Little is known about the profile of MFTD in postgraduate curricula for core and higher specialist training.

**Aims:** We profiled the inclusion of MFTD in the curricula of postgraduate core, higher and streamlined medical and surgical specialties in the Republic of Ireland and the UK. Methods: All publicly available syllabi of basic and higher/ streamlined specialist training in postgraduate medical and surgical colleges in both jurisdictions were analysed (N = 122).

**Results:** In Ireland, 25% of basic training schemes included MFTD in the curriculum. Two-thirds of curricula of higher specialty and streamlined training schemes also included MFTD. For the UK, 44% of core and 36% of higher training schemes curricula included MFTD. Just under one-quarter of all curricula reviewed included MFTD for more than one medical condition or treatment. Common topics in both Irish and UK curricula included seizures/epilepsy, syncope and visual disturbances.

**Conclusion:** There are notable deficits in MFTD training for specialists in Ireland and the UK. Common conditions which can significantly impair MFTD such as stroke, diabetes and alcohol use disorders are severely underrepresented and curricula should be revisited to include relevant training and guidance for MFTD for trainees.

**Main Messages:**

- Doctors have an important role in supporting safe driving among patients with a range of medical conditions
- Basic and higher specialist training curricula in a range of specialities in the British Isles are deficient in content relating to medical fitness to drive
- Curricular development for specialist training should include provision of concise and speciality-relevant guidance on medical fitness to drive

**Research Questions:**

- What barriers exist currently to the inclusion of medical fitness to drive and how could these be mitigated?
- What are the specialty-relevant prioritizations of medical conditions related to medical fitness to drive for inclusion in each curriculum?
- What resources and evidence are available to specialist training programmes to support the guidance for medical fitness to drive in their curricula?

## Introduction

The importance of mobility and transport as a vector of health and well-being is an increasingly recognised element of medical practice. Not only are there many interventions which support and enhance continued driving with relevant medical conditions (1) but also routine advice on such conditions is associated with a 45% reduction in crashes (2). There is concern that doctors are unaware of guidelines on supporting medical fitness to drive (MFTD) (3) (4) (5) (6).

Marshall et al. found that most specialists in Canada interviewed (68%) acknowledged that MFTD was an important part of their practice but only 33% felt confident in determining their patient’s MFTD (7). This indicates the large discrepancy between the importance of MFTD in modern medicine and the level of training provided for it.

When new guidelines on MFTD are introduced with a linked education programme, such as occurred in Ireland in 2014, the professional uptake can be substantial: 86% of general practitioners reported using the guidelines with 71% rating themselves as confident or very confident at assessing MFTD (8). This study also showed that GPs expressed interest in further training on MFTD. Case-based workshops and implementation of teaching programmes are probably the most effective format for improving physician evaluations of MFTD (9) (10).

Despite the importance of supporting patients and clinicians in driving decisions with relevant medical conditions, very few medical schools include MFTD as a specific element in their training courses (11). It is clearly also an important issue in postgraduate specialist training, however, no studies addressing how this features in postgraduate training were found as part of a literature search for this study. We therefore undertook a review of publicly available syllabi of basic and higher specialist training in a range of postgraduate medical and surgical colleges in Ireland and the UK so as to consider future strategy for planning appropriate MFTD inclusion in specialist training.

## Methods

Documents for curricula of all Republic of Ireland (ROI) and UK Core, Specialty and Streamlined medical training schemes were analysed for inclusion of education on MFTD. Documents were obtained from websites of the colleges between 11 and 23 March 2020. If the document was not available, the same websites were used to identify the scheme coordinator who was contacted for information regarding the curriculum. This amounted to 47 programs in Ireland and 75 programs in the UK. The curriculum obtained from each program was independently studied by each of two members of our research team. Each individual noted where they found reference to fitness to drive and what it related to. If education on MFTD was present, the key areas or modules where it was seen were noted including stroke, seizure (epilepsy), with syncope, with use of particular drugs and with visual disturbances. Both parties then combined their results: as there was concordance, there was no indication for referral for third party adjudication as planned in the protocol.

## Results

### Republic of Ireland

In postgraduate medical training in the ROI, of the basic specialty training schemes analysed (General Paediatrics, Histopathology, Obstetrics and Gynaecology (Obs&Gyn), General Internal Medicine), one provided training on MFTD (Obs&Gyn) in relation to the effects of pharmacology/ drugs on MFTD.

Of the 43 higher specialty and streamlined training schemes inspected, 29 (67%) provided training on MFTD (Table 1). Just over one-half, (15) of these curricula provided training on MFTD in more than one area. Thirteen of these taught MFTD in relation to the effects of pharmacology/ drugs with no further mention of MFTD in relation to any particular medical conditions. The most prominent MFTD topics included MFTD with pharmacology/drugs (25), seizure/epilepsy (11), visual disturbances (3), syncope (3), diabetes (3) and stroke (2). Unitary representation was noted for sleep disorders, FTD relating to psychiatry, driving accessibility for the disabled, following an amputation, and FTD in relation to general cardiac, respiratory health (such as with chronic obstructive pulmonary disease), carotid artery disease and blackouts/ drop attacks (Figure 1).

**Table 1:** Analysis of Higher Specialty and Streamlined Training (ROI)

| Speciality | Education | Stroke | Epilepsy/Seizures | Syncope | Pharmacology/Drugs | Visual | Other | No. of Areas |
| --- | --- | --- | --- | --- | --- | --- | --- | --- |
| Cardiology | Yes |  | Yes | Yes | Yes |  |  | 3 |
| Chemical Pathology | Yes |  |  |  | Yes |  | Diabetes | 2 |
| Clinical Microbiology | Yes |  |  |  | Yes |  |  | 1 |
| Clinical Pharm and Therapeutics | Yes |  | Yes |  | Yes |  |  | 2 |
| Clinical Genetics | Yes |  |  |  | Yes |  |  | 1 |
| Dermatology | Yes |  |  |  | Yes |  |  | 1 |
| Diagnostic Radiology | No |  |  |  |  |  |  | 0 |
| Endocrinology | Yes |  | Yes |  | Yes |  | Diabetes | 3 |
| Gastroenterology | Yes |  | Yes |  | Yes |  |  | 2 |
| Genito-Urinary Medicine | Yes |  |  |  | Yes |  |  | 1 |
| Geriatrics | Yes | Yes | Yes | Yes | Yes |  |  | 4 |
| Haematology | Yes |  |  |  | Yes |  |  | 1 |
| Histopathology | Yes |  |  |  | Yes |  |  | 1 |
| Immunology | Yes |  |  |  | Yes |  |  | 1 |
| Infectious Diseases | Yes |  | Yes |  | Yes |  |  | 2 |
| Medical Oncology | Yes |  |  |  | Yes |  |  | 1 |
| Neonatology | No |  |  |  |  |  |  | 0 |
| Nephrology | Yes |  | Yes |  | Yes |  |  | 2 |
| Neurology | Yes |  | Yes |  | Yes | Yes | Sleep Disorders | 4 |
| Neuropathology | Yes |  |  |  | Yes |  |  | 1 |
| Ob&Gyn | Yes |  |  |  | Yes |  |  | 1 |
| Occupational Medicine | Yes |  |  |  | Yes |  |  | 1 |
| Paediatrics | No |  |  |  |  |  |  | 0 |
| Palliative Medicine | Yes |  |  |  | Yes |  |  | 1 |
| Psychiatry | Yes |  |  |  |  |  | Application of FTD in Psychiatry | 1 |
| Public Health Medicine | Yes |  |  |  | Yes |  |  | 1 |
| Radiation Oncology | No |  |  |  |  |  |  | 0 |
| Rehabilitation Medicine | Yes |  |  |  | Yes | Yes | Driving Accessibility for the Disabled, Driving after Amputation | 4 |
| Respiratory Medicine | Yes |  |  |  | Yes |  | General Respiratory Health (COPD) | 2 |
| Rheumatology | Yes |  | Yes |  | Yes |  |  | 2 |

| **Specialty**<br>**(Streamlined)** | **Education** | **Stroke** | **Epilepsy/**<br>**Seizures** | **Syncope** | **Drugs** | **Vision** | **Other** | **No of areas** |
| --- | --- | --- | --- | --- | --- | --- | --- | --- |
| Anaesthesia | No |  |  |  |  |  |  | 0 |
| General Practice | Yes |  | Yes |  |  | Yes | General<br>Cardiovascular<br>Health, Diabetes | 4 |
| Cardiothoracic<br>Surgery | No |  |  |  |  |  |  | 0 |
| Otolaryngology | No |  |  |  |  |  |  | 0 |
| Plastic Surgery | No |  |  |  |  |  |  | 0 |
| General Surgery | Yes | Yes |  |  |  |  | Carotid Artery<br>Disease | 2 |
| Urology | No |  |  |  |  |  |  | 0 |
| Trauma &<br>Orthopaedic Surgery | No |  |  |  |  |  |  | 0 |
| Neurosurgery | No |  |  |  |  |  |  | 0 |
| Paediatric Surgery | No |  |  |  |  |  |  | 0 |
| Medical<br>Ophthalmology | No |  |  |  |  |  |  | 0 |
| Surgical<br>Ophthalmology | No |  |  |  |  |  |  | 0 |
| Emergency Medicine | Yes |  | Yes | Yes |  |  | Blackout/ Drop<br>Attacks | 3 |

**Figure 1:**
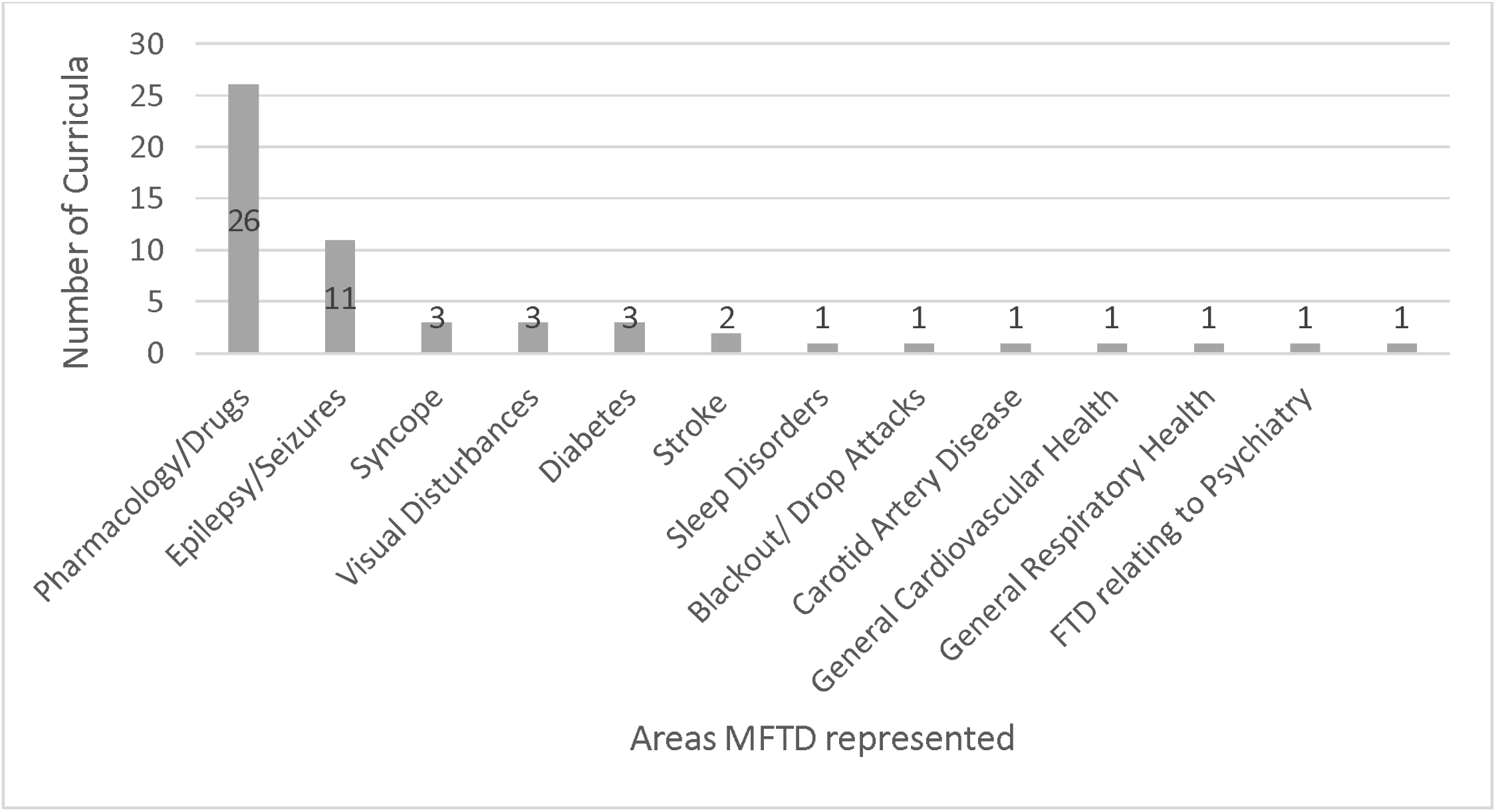
Education on MFTD in relation to specific areas (ROI) (Basic, Higher Specialty and Streamlined Training)

### United Kingdom

Of 9 core training curricula, 4 mentioned MFTD (44%) (Table 2). Key areas of MFTD represented included epilepsy/seizures (3), syncope (3), blackout/drop attack (3), general legislation (2), visual disturbances (2) and MFTD following surgery (1). Three provided education on MFTD in more than one area.

**Table 2:** Analysis of Core Training Curricula (UK)

| Specialty | Education | Legislation | Stroke | Epilepsy/<br>Seizures | Syncope | Blackouts | Vision | Other | No. of<br>Areas |
| --- | --- | --- | --- | --- | --- | --- | --- | --- | --- |
| Acute care common stem | Yes | Yes |  | Yes | Yes | Yes | Yes |  | 5 |
| Core medical training | Yes | Yes |  | Yes | Yes | Yes | Yes |  | 5 |
| Core anaesthetics training | Yes |  |  |  |  |  |  | Post-surgery | 1 |
| Core surgical training | No |  |  |  |  |  |  |  | 0 |
| Core psychiatry | No |  |  |  |  |  |  |  | 0 |
| Core radiology training | No |  |  |  |  |  |  |  | 0 |
| Broad based training | Yes |  |  | Yes | Yes | Yes |  |  | 3 |
| Core Ob & Gyn Training | No |  |  |  |  |  |  |  | 0 |
| Paediatrics Core Training | No |  |  |  |  |  |  |  | 0 |

Of 66 higher specialty curricula analysed (Table 3), one-third, (24, 36%) included MFTD while 42 (64%) did not. Of the 24 curricula which included MFTD, 13 mentioned MFTD in terms of general legislation with no further mention of MFTD in relation to any particular medical conditions. The most common specific conditions where MFTD was represented were syncope (6), visual disturbances (6), seizures/epilepsy (5), blackouts/drop attacks (5) and diabetes (2) (Figure 2). MFTD was noted in single instances for stroke, vertigo, general cardiovascular health including arrhythmias and pacemakers, carotid artery disease, sleep disorders, pregnancy, pharmacology/drugs and general respiratory health (Figure 2). Of the 24 curricula which provided MFTD, only 10 (42%) provided education in more than one area.

**Table 3:**
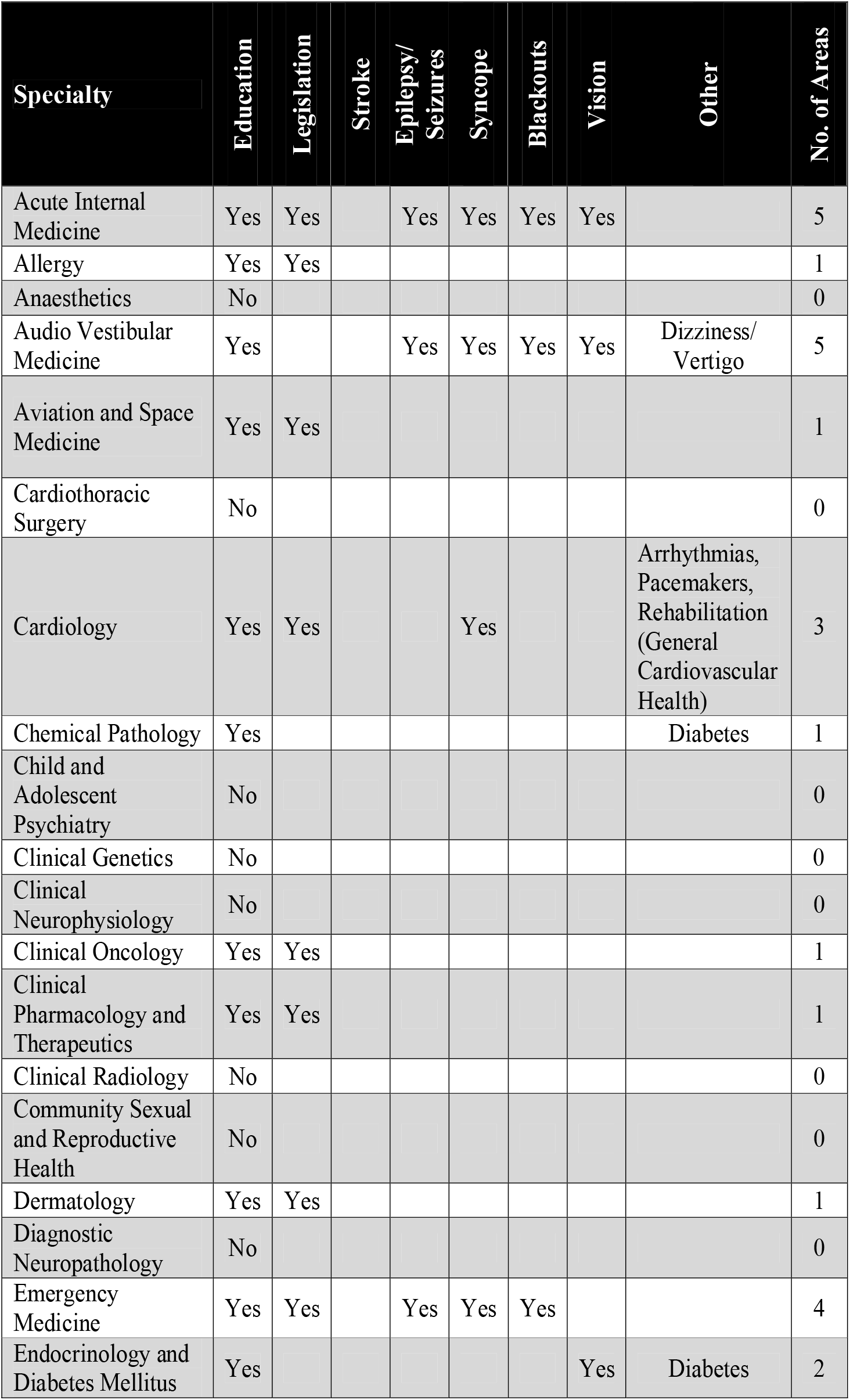
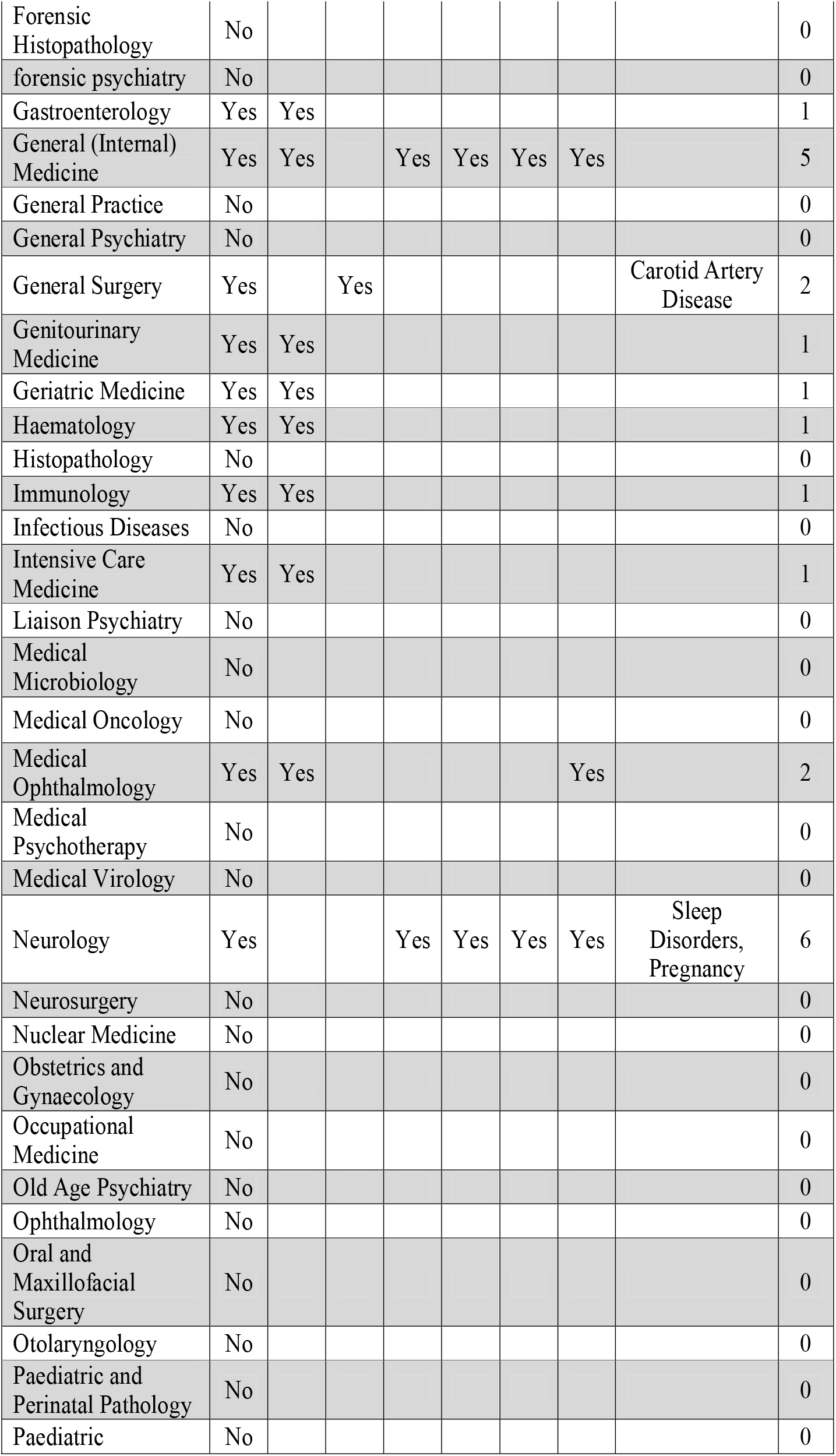
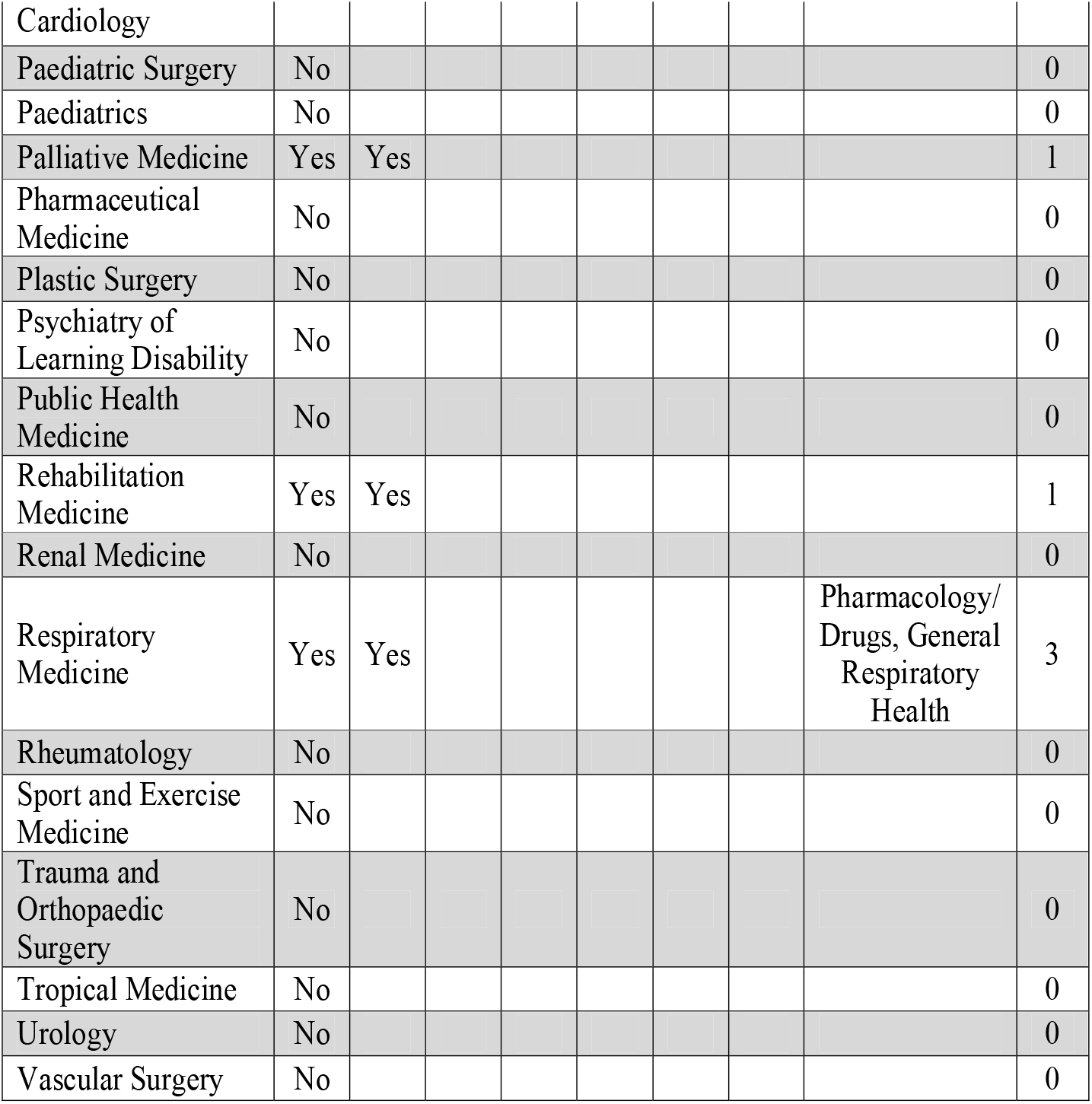
Analysis of Higher Specialist and Streamlined Curricula (UK)

**Figure 2:**
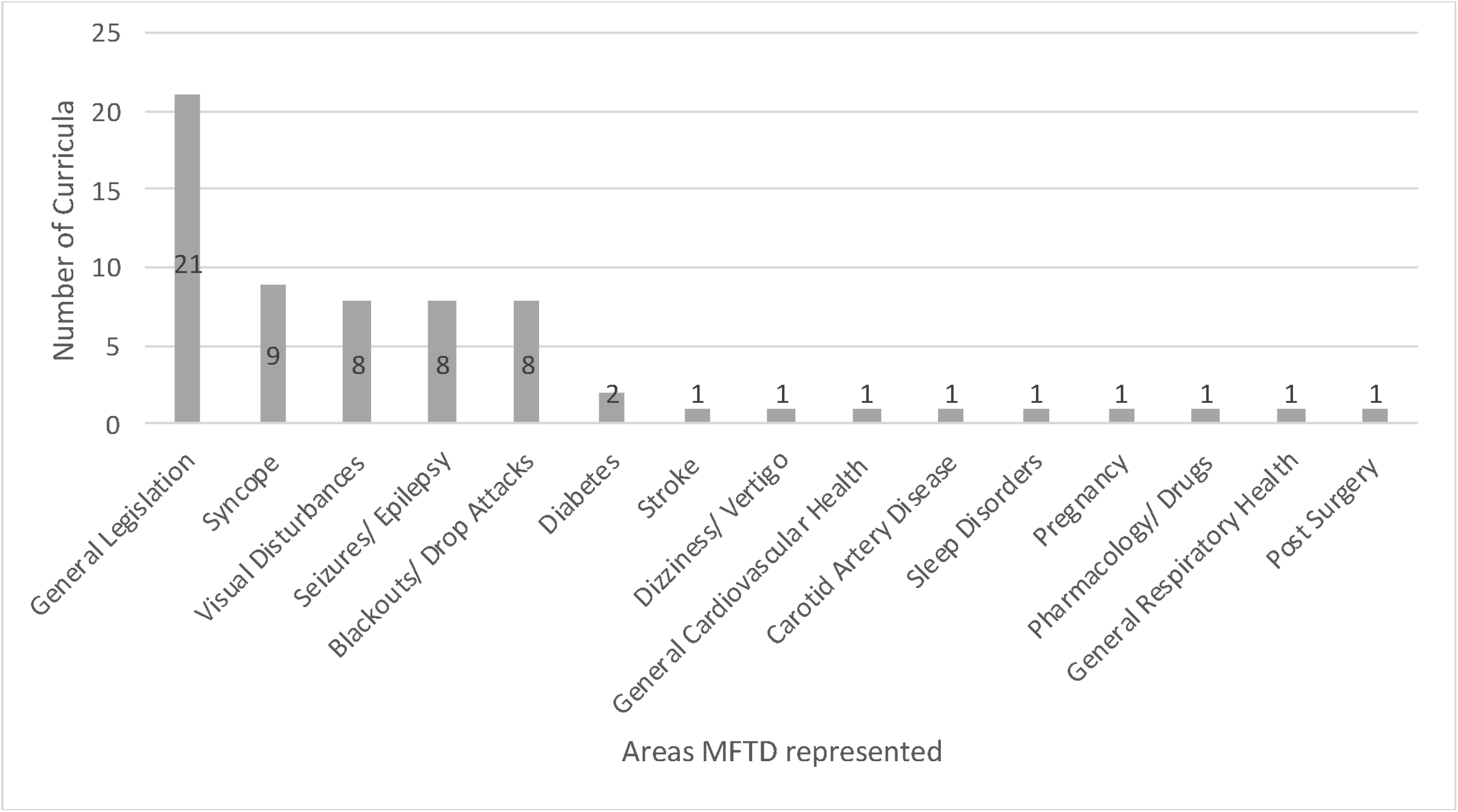
Education on MFTD in relation to specific areas (UK) (Core and Higher Training)

When data from Ireland and the UK are merged, 5 of 13 (38%) core/basic specialty curricula provided some level of exposure to MFTD and 53 of the 109 (49%) higher specialty/streamlined curricula included MFTD. Just under one quarter of all curricula analysed provided training on MFTD in more than one area (23%) (Figure 3).

**Figure 3:**
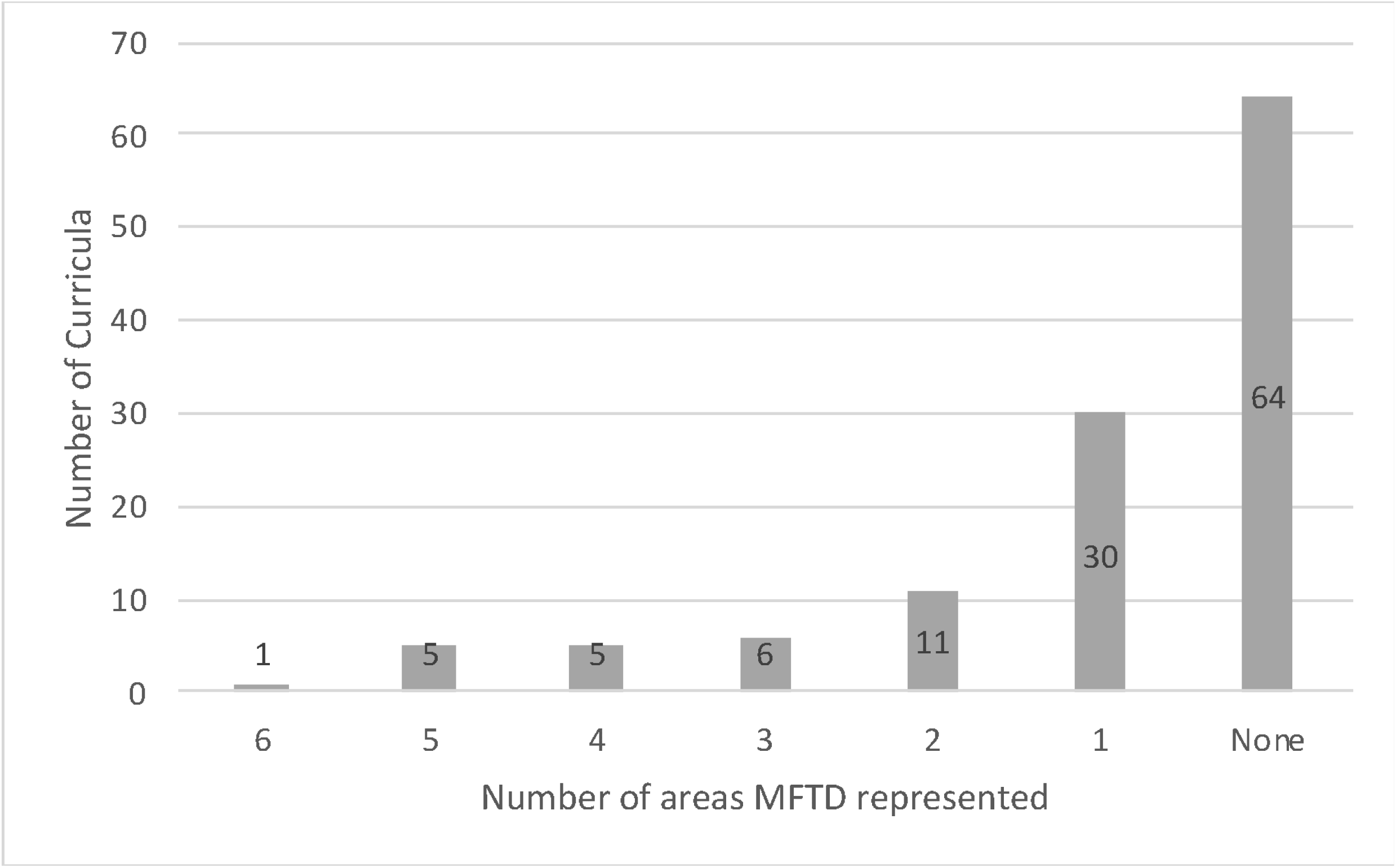
Number of areas MFTD represented (All Curricula) (UK and ROI)

Most commonly represented areas (both core and higher training) included seizures/epilepsy (19), syncope (12), visual disturbances (11) and diabetes (5). No training scheme mentioned alcohol use disorders related to MFTD. There was no mention of MFTD related to medicine for older people or neurodegenerative conditions such as multiple sclerosis or Parkinson’s Disease.

## Discussion

Our research indicates a significant deficit and disequilibrium in subject matter in the training of specialists in Ireland and in the UK in learning to support patients with issues related to MFTD. We are mindful that different specialties will have different profiles of patient morbidity, and therefore of relative salience of particular aspects of MFTD to their clinical practice. However, we also practice in an environment where co-morbidity is increasingly present in our patient populations, and there is a broad range of conditions applicable across medical specialties. Many of these are conditions that affect MFTD and are underrepresented in the postgraduate training curricula. Any review of developing MFTD in postgraduate training is an opportunity to evaluate which range of conditions should feature on each curriculum.

Across both Ireland and the UK, only 38% of core/basic and 49% of higher specialty/streamlined curricula provided any training on MFTD. Further analysis indicates that of all curricula analysed, only 23% provided training in more than one area, highlighting significant deficits in MFTD training to support patients.

The coverage of MFTD generally follows the historical emergence of conditions in guidelines, with seizures and syncope as clearly tangible impediments to safe driving (12). Epilepsy/seizures was one of the more frequently included areas in the programmes reviewed, reflecting the early prominence of epilepsy in MFTD regulations since the 1920s (12). Epilepsy is a common condition across various specialties and it is estimated to affect 1% of the Irish population (13). Up to 20% of patients on treatment continue to experience seizures which can involve loss of consciousness (and/ or motor control) and this can severely impair the ability to drive (14). This has obvious implications for MFTD and is a topic that needs to be included in more training curricula.

In a similar manner, MFTD related to syncope also featured relatively frequently. Syncope is a common condition with a frequency of 15-39% (15) and can significantly impair MFTD (16) due to sudden loss of consciousness. Syncope is a condition which will most likely be seen by most doctors throughout their careers and its important effects on MFTD should be on more curricula.

The most notable deficit was that related to alcohol use disorders and other substance use disorders. Not only is alcohol use disorder a common co-morbidity across all adult groups encountered in virtually all clinical practice but also a significant factor in causation of fatal and injurious traffic crashes: drivers with alcohol abuse problems encounter accidents more frequently than those without (17). The absence of advice almost certainly reflects a wider ambivalence among doctors about engaging with diagnosis and advice for their patients with alcohol use disorders (18). This is of particular concern as a number of studies have indicated that brief counselling of those affected by alcohol use disorders admitted to general wards (19) and primary care is effective in reducing drinking and deaths (20).

Other common conditions not featuring adequately include visual impairment, a common presentation in medicine: conditions such as cataracts can considerably affect MFTD (21). The overall occurrence of guidance on visual impairments on postgraduate training schemes was low and did not acknowledge how commonly it is encountered in clinical practice. As many branches of medicine will engage with patients who have had stroke, the profile of MFTD relating to stroke was also underrepresented (3 curricula), despite evidence that a large proportion of stroke survivors have challenges with returning to drive (22). Many of these are also left to make decisions on their MFTD without professional advice, indicating likely deficits in physician awareness of guidelines (23).

Although not as underrepresented as stroke, there was a lack of curricular presence regarding MFTD with diabetes mellitus, also commonly encountered across a wide variety of specialties. Although associated with a very modest increased relative risk of motor vehicle crash compared to non-diabetic controls (24), it is important that those affected are given appropriate advice on continued safe driving.

A further notable absence was MFTD training for neurodegenerative diseases such as multiple sclerosis (25) and Parkinson’s Disease (PD): drivers with PD have been shown to drive in a less safe manner on the road compared to healthy controls (26).

Given that older drivers are the group most likely to present with conditions relevant to MFTD and who can benefit from a range of interventions to support continued driving (1), there was also no indication of MFTD for older populations in the postgraduate training schemes in Ireland and the UK. The opportunities for curriculum development for older drivers is exemplified by a course delivered to healthcare professionals in California between 2009 and 2011: afterwards 92% of participants reported a greater understanding of MFTD and how to advise their older patients about driving(27).

Previous research has already shown that doctors are largely unaware of guidelines supporting MFTD (3) (4) (5) (6). This may in part relate to the findings of our study that that most trainees in the British Isles do not receive an adequate level of training regarding MFTD at either core or higher specialty medical training levels. We would recommend that education on MFTD be included routinely in all postgraduate medical curricula with an appropriate emphasis on rehabilitation and enabling continued driving, barriers to continued safe driving (28) and alternative transport options for those who can no longer drive. This might usefully incorporate in core/basic specialist training the principles of assessment and management for common conditions, as well as familiarization with national guidelines, driving assessment resources and relevant legislation. Aspects relating more clearly to conditions more closely associated with each higher specialization could be further developed during higher specialist training. Being supported in continued safe driving is an important goal for our patients (29), and it is important that our professional training should reflect our opportunity to support this most central of activities for those we serve.

## Data Availability

Available on request and in tables in paper

